# Blood biomarker changes and relationships after low dose oral ketamine treatment for post-traumatic stress disorder (PTSD)

**DOI:** 10.1101/2025.03.02.25323193

**Authors:** Bonnie L. Quigley, Emerald Orr, Sophie Kafka, Maryam Hajishafiee, Ana P. Bouças, Nathan Wellington, Megan Dutton, Monique Jones, Fiona Randall, Jim Lagopoulos, Adem T. Can, Daniel F. Hermens

## Abstract

Ketamine has been investigated as a treatment alternative for PTSD for the last 20 years, yet there have been virtually no reports of biological changes or biomarker characterisation related to treatment. To address this significant gap, this study analysed blood samples from 25 participants with PTSD who took part in an open-label 6-week trial of low dose oral ketamine treatment. Serum and plasma samples were quantified before and after ketamine treatment for brain-derived neurotrophic factor (BDNF), vascular endothelial growth factor A (VEGF-A), serotonin, FK506 binding protein 51 (FKBP51) and a panel of cytokines (interleukin (IL)-1β, IL-2, IL-4, IL-6, IL-12p70, IL-17A and tumour necrosis factor alpha (TNFα)). Analysis of BDNF and VEGF-A levels detected a significant positive correlation between the two biomarkers and a small but statistically significant decrease in both measures after ketamine treatment. This novel finding reinforces evidence that ketamine’s effects may rely on a reciprocal interaction between BDNF and VEGF-A, offering potential insights into a biological mechanism underpinning PTSD symptom reduction. Additionally, the analysis of FKBP51 and serotonin revealed novel relationships between these biomarkers and clinical scales, before and after ketamine treatment. Finally, significant changes or relationships involving the immune cytokines were not detected, possibly because half the participants presented with low-grade inflammation while the other half did not. This study represents the first comprehensive analysis of blood biomarkers before and after ketamine treatment for PTSD and reveals important biological changes and relationships related to this treatment.

## Introduction

Trauma is relatively common, with a consensus indicating more than 70% of people will experience a traumatic event in their lifetime^1^. From these experiences, ∼10% of people will go on to develop post-traumatic stress disorder (PTSD), characterised by re-experiencing, avoidance, increased negative affect and hyperarousal symptoms^2, 3^. In contemporary clinical settings, PTSD treatment often includes a combination of psychological and pharmacological interventions^4, 5^. Exposure-based psychotherapies have the largest evidence base and aim to reduce symptoms by encouraging adaptive associations with painful memories, rendering the trauma tolerable^6^. However, treatment response rates from current exposure-based protocols report only 46-60% of patients achieve recovery with these treatments^7^. Antidepressants, particularly selective serotonin reuptake inhibitors (SSRIs), are the primary treatment medication for PTSD in pharmacotherapy^8^. Two SSRIs are FDA-approved for PTSD treatment, paroxetine and sertraline, and response rates to these drugs rarely exceed 60%, with less than 30% of patients achieving full remission^8, 9^.

Ketamine, an N-methyl-D-aspartate-type (NMDA) receptor antagonist, has been investigated as a treatment for both treatment-resistant depression (TRD) and PTSD with promising results^10, 11^. To date, investigations on ketamine for PTSD have primarily used intravenous (IV) infusion. Recently, our group completed an open-label clinical trial using low dose oral ketamine for PTSD symptom reduction (one dose weekly for six weeks in a titrating up manner), with a treatment response rate of 73% one-week post-treatment and 59% one-month post-treatment^12^. The response rate to oral ketamine was comparable to those reported in IV ketamine PTSD trials^11^. However, oral ketamine offers the advantages of being more practical for use in conventional clinical psychiatry settings and significantly more cost-effective than other administration methods^13-15^.

However, a significant gap in our understanding of ketamine as a treatment for PTSD lies in identifying the biological changes associated with its effects. The investigation of biomarkers associated with ketamine treatment in humans has been notably absent from the literature, even in the more extensively studied field of ketamine for TRD^16^. Recent reviews summarizing blood biomarkers related to ketamine and TRD identify only eight studies on to the topic, primarily focusing on brain-derived neurotrophic factor (BDNF), serotonin, interleukin (IL)-6, S100β and various metabolomic biomarkers^17, 18^. Given the 25-year history of ketamine research in TRD, this scarcity is remarkable. Research into ketamine’s potential for PTSD treatment began 20 years ago^11^, yet the only reported blood biomarker reported in this area is d-serine, with higher plasma levels linked to shorter remission times in a ketamine-psychotherapy trial^19^. The remainder of the PTSD biomarker literature focuses on biomarker differences between cohorts with and without PTSD or biomarkers that confer risk of developing PTSD^20^.

Important biomarkers that have been investigated in relation to PTSD include circulating BDNF, where meta-analyses show an overall *increase* in BDNF during the clinical condition of PTSD, opposite to every other psychiatric disorder investigated^21^. Additionally, immune cytokines often present as a systemic, low-grade inflammation in PTSD patients^22, 23^. Other potential PTSD biomarkers with less extensive literature include serotonin^24^, where PTSD cohorts have been reported as having lower circulating levels and FK506 binding protein 51 (FKBP51)^25^, an important stress response co-chaperone with genetic polymorphisms linked to PTSD development^26^. Finally, given the connection between BDNF and vascular endothelial growth factor A (VEGF-A) in antidepressant treatment response^27, 28^, our group recently determined that a cohort of older adults with PTSD had higher levels of VEGF-A compared to non-PTSD matched controls^29^. Given the lack of specific biological investigation related to treatment, a critical first step in understanding how ketamine treatment may lead to the clinical improvements in PTSD is to determine what is happening to these common biomarkers before and after treatment.

To address this critical gap in the literature on the biological effects of ketamine in PTSD treatment, we analysed circulating levels of BDNF, VEGF-A, serotonin, FKBP51 and a panel of cytokines (IL-1β, IL-2, IL-4, IL-6, IL-12p70, IL-17A and tumour necrosis factor alpha (TNFα)) before and after low dose oral ketamine treatment. This investigation aimed to identify potential biomarker changes and their relationships to PTSD symptomology and ketamine administration.

## Materials and Methods

### Study participants

Samples for this analysis were collected from the Oral Ketamine Trial on PTSD (OKTOP), with the full details and primary outcomes of the open-label clinical trial described in Quigley et al.^12^. The trial was approved by the Metro North Health Human Research Ethics Committee (HREC) (Project ID: 42836; HREC/18/QPCH/28), the University of the Sunshine Coast HREC (A181190) and prospectively registered with the Clinical Trials Registry of Australia and New Zealand (ACTRN12618001965291). To maximize the number of samples for analysis, trial participants diagnosed with PTSD via the Clinician Administered PTSD Scale for DSM-5 (CAPS-5) were included in this study if blood samples were available from the baseline assessment before ketamine treatment (“pre-treatment”) and at least one sample after ketamine treatment (“post-treatment” - ∼2 hours after the sixth/final ketamine dose, “follow-up 1-week” – 1-7 days after the sixth/final ketamine dose and/or “follow-up 1-month” – 28-32 days after the sixth/final ketamine dose). This generated a study cohort of n=25 participants (**Table 1**), consisting of n=25 pre-treatment samples, n=19 post-treatment samples, n=24 follow-up 1-week samples and n=23 follow-up 1-month samples. Additionally, n=14 mid-treatment samples (∼2 hours after the third ketamine dose) were included in the biomarker change analysis only. Response to ketamine treatment was defined in the trial as ≥50% reduction in PTSD Checklist for DSM-5 (PCL-5) score from pre-treatment^12^, with these samples representing response rates of # responders/# non-responders/# missing samples or outcomes of 15/3/7 at post-treatment, 15/8/2 at follow-up 1-week and 12/11/2 at follow-up 1-month for all 25 participants, respectively.

**Table 1.** Participant demographics

| Characteristic | Value* |
| --- | --- |
| # Participants | n = 25 |
| Sex | 16F / 9M |
| Age | 47.8 ± 13.0 (22 - 77) |
| PTSD duration (years) | 18.8 ± 16.2 (2 - 55) |
| CAPS-5 score | 54.2 ± 7.0 (41 - 67) |
| Index trauma |  |
| Childhood trauma | 11 (44%) |
| Military/war | 4 (16%) |
| Sexual assault/abuse and/or DV | 4 (16%) |
| Physical assault/abuse/accident | 4 (16%) |
| Emergency services (work) | 1 (4%) |
| Bereavement (death of child) | 1 (4%) |
| Highest education |  |
| Secondary year 10 | 1 (4%) |
| Secondary year 12 | 4 (16%) |
| Tertiary | 12 (48%) |
| Post-graduate | 6 (24%) |
| Unknown | 2 (8%) |
| Major Depressive Disorder (MDD) | 19 (76%) |
| Complex PTSD (cPTSD) | 3 (12%) |
| Psychotropic medications |  |
| Benzodiazepines | 13 (52%) |
| SSRIs | 4 (16%) |
| SNRIs | 6 (24%) |
| Tetracyclic antidepressants | 3 (12%) |
| Others | 9 (40%) |

\* Mean ± standard deviation (range) or count (percentage), as appropriate. CAPS-5 = Clinician-Administered PTSD Scale for DSM-5. DV = domestic violence. SSRI = Selective serotonin reuptake inhibitors. SNRI = Serotonin and norepinephrine reuptake inhibitors.

### Clinical measures

In addition to the CAPS-5^30^ conducted at pre-treatment to confirm study eligibility, the following clinical rating scales were completed by participants at pre-treatment, post-treatment and follow-up timepoints to assess study outcomes: (a) PCL-5, a 20-item self-report measure often used in conjunction with the CAPS-5 to monitor PTSD symptom change across patient treatment^30, 31^; (b) Depression, Anxiety, and Stress Scale (DASS-21), 21-item self-report measure assessing perceived depressive, anxiety, and stress symptoms^32^; and (c) World Health Organization Well-Being Index (WHO-5), a 5-item self-report global rating scale measuring subjective well-being^33^.

### Sample collection and processing

Non-fasting whole blood was collected at pre-treatment (week 0), mid-treatment (week 3), post-treatment (week 6), follow-up 1-week (week 7) and follow-up 1-month (week 10) timepoints at midday/early afternoon by a certified phlebotomist using serum separator tubes (SST) and K2EDTA (plasma) collection tubes. After a minimum 30 minutes at room temperature, an aliquot of K2EDTA whole blood was removed and stored at -80°C before all tubes were centrifuged at 2465 x g for 15 min at 4°C. All samples were processed within four hours of collection. Serum and plasma were removed from their respective tubes, aliquoted and stored at -80°C.

### Biomarker quantification

Before analysis, all samples were thawed on ice and centrifuged at 10,000 x g for 10 min at 4°C. BDNF, VEGF-A and serotonin were quantified from serum while FKBP51 and all cytokines were quantified from plasma. Standard ProcartaPlex simplex assays (ThermoFisher Scientific, Australia) were combined for the detection of BDNF (lower limit of quantification (LLOQ) of 2.03 pg/ml) and VEGF-A (LLOQ of 4.88 pg/ml) while high-sensitivity assays were used to quantify IL-1β (LLOQ of 0.28 pg/ml), IL-2 (LLOQ of 0.88 pg/ml), IL-4 (LLOQ of 1.29 pg/ml), IL-6 (LLOQ of 1.29 pg/ml), IL-12p70 (LLOQ of 0.77 pg/ml), IL-17A (LLOQ of 0.30 pg/ml) and TNFα (LLOQ of 0.62 pg/ml). Assays were conducted according to the manufacturer’s instruction, read on a Luminex 200 instrument (ThermoFisher) and processed using the ProcartaPlex Analysis App (ThermoFisher). Enzyme-linked immunosorbent assays (ELISAs) were used to quantify serotonin (Fast Track kit, Immusmol, France; LLOQ of 15 ng/ml) and FKBP51 (Human kit, Invitrogen, ThermoFisher, Australia; LLOQ of 0.41 ng/ml) according to the manufacturer’s instructions and processed using GainData (https://www.arigobio.com/elisa-analysis). BDNF, VEGF-A and serotonin levels were within assay detectable ranges for all samples tested while detectable cytokine levels ranged from 95% (IL-1β and IL-12p70), 90% (IL-4 and IL-6), 80% (IL-2) and 72% (IL-17A and TNFα) of samples tested. FKBP51 levels were within the assay detectable range for 66% of samples. Samples with targets below their assay’s LLOQ were assigned a value of half their LLOQ for statistical analysis.

### Statistical analysis

All analyses were performed using R Statistical Software (v4.3.3)^34^. Overall changes in biomarker levels by study timepoint were calculated using the Kruskal-Wallis test. Changes in biomarker levels over the course of the ketamine trial were assessed by linear mixed model analysis using lme4^35^ and lmerTest^36^ and visualised using ggplot2^37^. Models used change values for each biomarker, which were calculated by subtracting log transformed pre-treatment levels from log transformed subsequent timepoint levels (similar to ^38^). The cytokine heatmap was generated with heatmap.2 (in gplots)^39^ after scaling samples within each target (subtract target mean and divide by the target standard deviation). Comparisons of biomarker levels by immune group were calculated using the Wilcoxon rank sum test, with p-values adjusted for multiple comparisons using Benjamini-Hochberg (BH) false discovery rate method^40^. Spearman rank correlations were calculated using rcorr (in Hmisc)^41^, visualised using corrplot^42^ and p-values adjusted using the BH false discovery rate method.

## Results

### Biomarker changes throughout ketamine treatment and follow-up

Average quantities for each biomarker tested are reported in **Table 2** by timepoint. In terms of means, no biomarker levels were found to significantly change throughout the trial (by Kruskal-Wallis test, **Table 2**).

**Table 2.** Mean blood biomarker levels throughout ketamine treatment trial

| Marker | Pre-treatment<br>(n=25) | Mid-treatment<br>(n=14) | Post-treatment<br>(n=19) | Follow-up 1<br>week (n=24) | Follow-up 1<br>month (n=23) | Kruskal-Wallis test |
| --- | --- | --- | --- | --- | --- | --- |
| BDNF (pg/ml) | 261.34 ± 347.78 | 205.12 ± 294.41 | 170.94 ± 225.61 | 219.80 ± 285.80 | 219.80 ± 285.80 | H(4) = 2.06, P = 0.725 |
| VEGF-A (pg/ml) | 323.62 ± 269.86 | 287.21 ± 272.13 | 309.45 ± 225.03 | 295.71 ± 262.27 | 302.61 ± 303.85 | H(4) = 1.01, P = 0.908 |
| Serotonin (ng/ml) | 89.95 ± 75.30 | ND | 105.11 ± 106.96 | 83.82 ± 83.17 | 93.72 ± 80.78 | H(3) = 0.59, P = 0.899 |
| FKBP51 (ng/ml)* | 20.92 ± 59.62 | ND | 10.60 ± 28.25 | 16.99 ± 46.45 | 17.55 ± 46.75 | H(3) = 0.06, P = 0.996 |
| IL-1β (pg/ml)* | 1.88 ± 1.34 | 2.24 ± 1.89 | 2.27 ± 1.91 | 2.91 ± 2.42 | 2.70 ± 1.82 | H(4) = 3.70, P = 0.448 |
| IL-2 (pg/ml)* | 2.24 ± 3.03 | 3.69 ± 4.42 | 3.17 ± 4.12 | 4.01 ± 3.63 | 3.99 ± 3.51 | H(4) = 7.38, P = 0.117 |
| IL-4 (pg/ml)* | 1.91 ± 1.14 | 2.81 ± 2.39 | 2.07 ± 1.43 | 3.16 ± 2.53 | 2.95 ± 1.79 | H(4) = 6.26, P = 0.181 |
| IL-6 (pg/ml)* | 4.01 ± 6.47 | 6.14 ± 11.02 | 4.62 ± 8.59 | 4.42 ± 7.03 | 5.81 ± 11.81 | H(4) = 3.65, P = 0.455 |
| IL-12p70 (pg/ml)* | 6.49 ± 4.83 | 6.30 ± 5.63 | 4.63 ± 3.31 | 6.46 ± 5.97 | 5.94 ± 3.70 | H(4) = 2.00, P = 0.735 |
| IL-17A (pg/ml)* | 0.79 ± 0.66 | 0.88 ± 0.97 | 0.61 ± 0.66 | 1.00 ± 1.12 | 0.88 ± 0.69 | H(4) = 3.40, P = 0.493 |
| TNFα (pg/ml)* | 3.19 ± 4.23 | 2.92 ± 2.76 | 2.72 ± 2.53 | 3.11 ± 2.93 | 2.70 ± 2.39 | H(4) = 0.17, P = 0.997 |

Values given as mean ± standard deviation. ND = Not determined. Asterisks denote targets where calculations include inferred quantities based on assay detection limits (details in Methods).

However, significant variation within each biomarker was observed, as evident by the large standard deviations in **Table 2**. To address this individual variability, additional data transformation and analysis was performed with the serum biomarkers BDNF, VEGF-A, and serotonin (where quantifiable results were obtained from all the samples tested (see methods)). Each participant’s BDNF, VEGF-A and serotonin quantities were log-transformed and subtracted from their pre-treatment values, thereby setting each participant’s pre-treatment value to “zero” and converting subsequent timepoint values to the change in biomarker level from pre-treatment. Linear mixed model analysis was then used to assess the pattern of change. Models considered change in biomarker quantity and study timepoints as fixed effects and individual participants as random effects. The age and sex of the study participants were tested as additional fixed effects and found not to be significant for these biomarkers in this dataset, so were omitted from the final models.

From this change analysis, BDNF and VEGF-A levels were found to show small but significant decreases after ketamine treatment (**Figure 1**). Small BDNF decreases from pre-treatment were detected at mid-treatment (decrease of 0.216 log10 pg/ml, p = 0.010), follow-up 1-week (decrease of 0.157 log10 pg/ml, p = 0.026) and follow-up 1-month (decrease of 0.142 log10 pg/ml, p = 0.046) (**Figure 1A**). Concurrently, significant decreases in VEGF-A levels from pre-treatment were detected at mid-treatment (decrease of 0.062 log10 pg/ml, p = 0.030), post-treatment (decrease of 0.052 log10 pg/ml, p = 0.043), follow-up 1-week (decrease of 0.065 log10 pg/ml, p = 0.007) and follow-up 1-month (decrease of 0.050 log10 pg/ml, p = 0.038) (**Figure 1B**). Serotonin levels did not show any significant change over the course of ketamine treatment.

**Figure 1.**
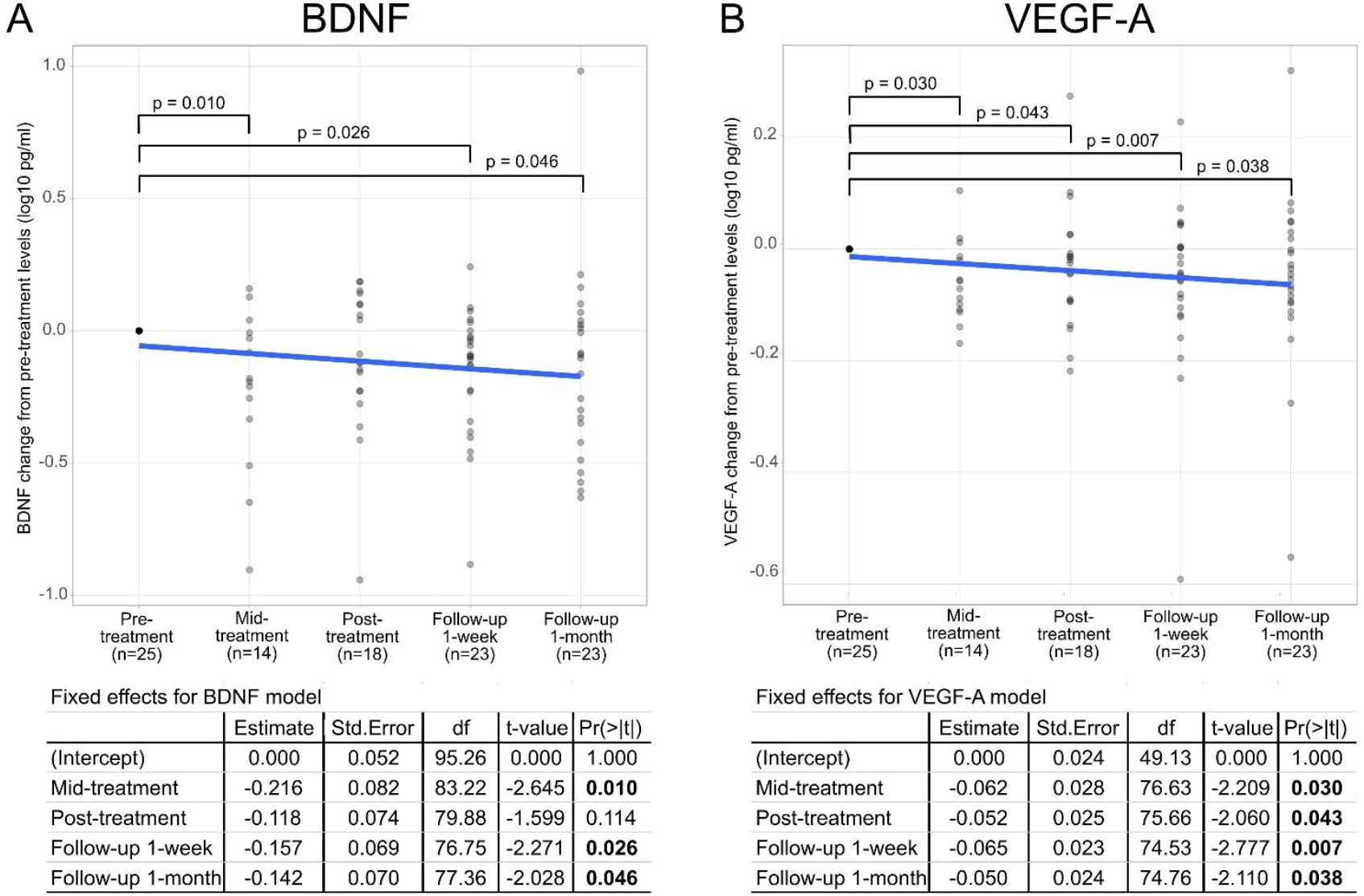
Linear mixed models (LMMs) for BDNF (A) and VEGF-A (B) levels throughout the trial. Change values for each biomarker were calculated by subtracting log10 pre-treatment values from each log10 timepoint value for each participant. The overall data trend is represented by the blue line and significance differences (p < 0.05) compared to pre-treatment are indicated. The fixed effect statistics for each model are given as a table under their respective graphs.

### Biomarker relationships by inflammatory status of participants

To understand the immune status of the study group better and investigate cytokine measurements in more detail, immune biomarkers were normalised and clustered for all study participants and timepoints (**Figure 2A**). This clustering revealed two distinct inflammatory groups within the study cohort: a “high” inflammatory group (n=12), with average cytokine levels above the group average (green colours in **Figure 2A**) and a “low” inflammatory group (n=13), with average cytokine levels below the group average (red colours in **Figure 2A**). Between these inflammatory groups, there was a statistically significant difference between IL-1β, IL-2, IL-4, IL-12p70, IL-17A and TNFα levels at all four major study timepoints (pre-treatment, post-treatment, follow-up 1-week and follow-up 1-month), with IL-6 having significant differences at pre-treatment and follow-up 1-week (**Supplementary Table 1**).

**Figure 2.**
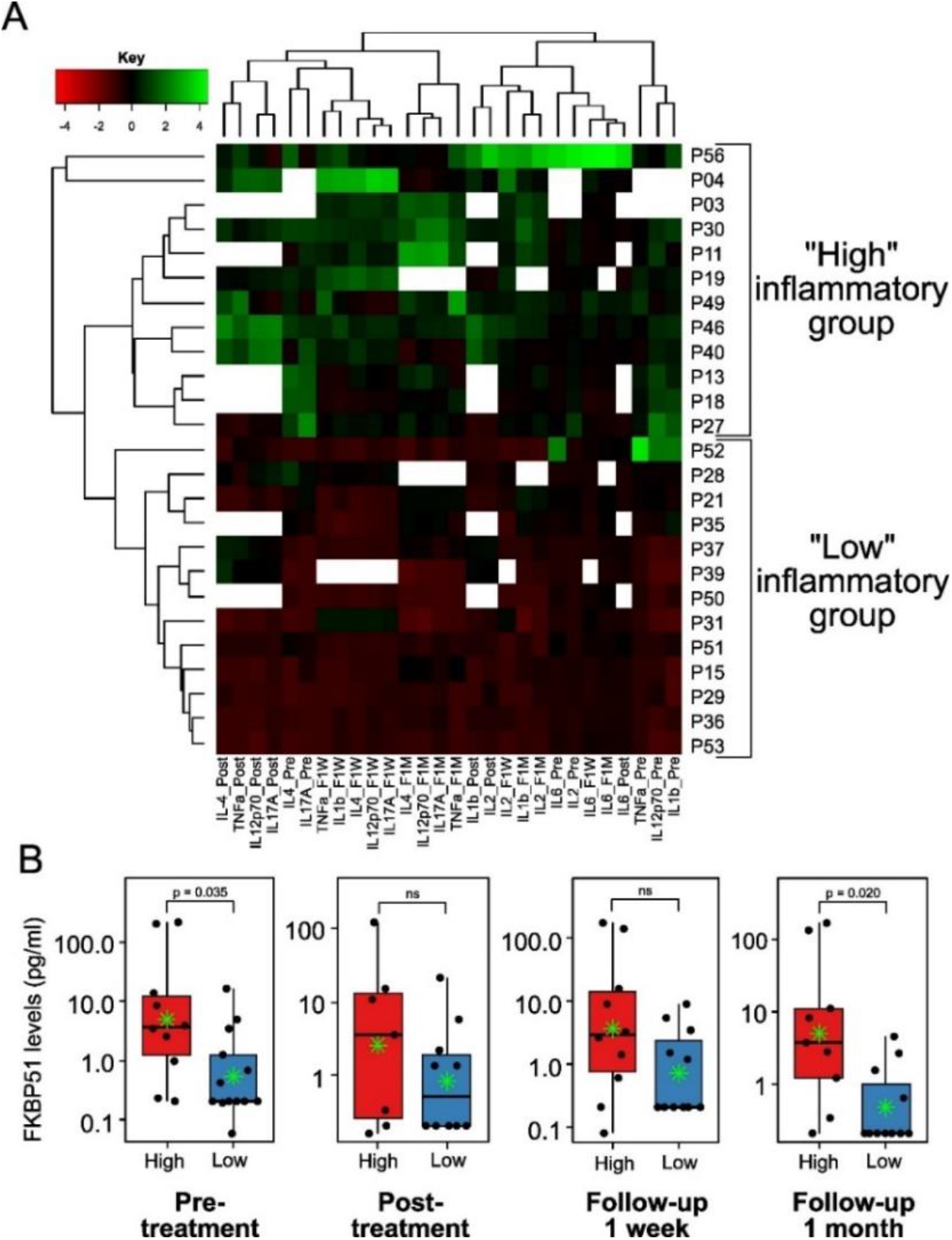
(A) Clustering of cytokine levels by participant and study timepoint. P# indicate individual participants (rows). Values for each cytokine at each timepoint (columns) were normalised for comparison (subtract the group mean and divide by the group standard deviation – generates black for average values, green for values above the average and red for values below the average). White squares indicate missing samples. _Pre = pre-treatment sample, _Post = post-treatment sample, _F1W = follow-up 1 week sample, _F1M = follow-up 1 month sample. (B) Box and whisker plots of FKBP51 values by inflammatory group status at study timepoints (note: mid-treatment samples were not tested for FKBP51, see Table 2). Green star denotes group mean. Wilcoxon rank sum test was used to determine statistical differences, with p-values adjusted for multiple comparisons using BH false discovery rate. ns = not significant (p > 0.05).

Examination of the inflammatory clusters revealed two important findings. The first was that ketamine did not appear to significantly alter circulating cytokine levels during or after treatment. The levels of each cytokine remained relatively consistent between timepoints (**Supplementary table 1**) and the distinct separation between the inflammatory groups was maintained throughout the treatment course (**Figure 2A**). There was no significant correlation or difference detected between a participant’s inflammatory group status and their personal demographics (sex, age, PTSD severity, and presence of co-morbid depression) or their treatment response in the trial, leaving the reason for the two distinct inflammatory subgroups unknown.

The results for FKBP51, on the other hand, revealed a significant association between FKBP51 levels and inflammatory group status (**Figure 2B**). At the four study timepoints where FKBP51 was tested, average FKBP51 levels were consistently higher in the “high” inflammatory group, with this difference reaching statistical significance at the pre-treatment and follow-up 1-month timepoints (**Figure 2B, Supplementary table 1**). No difference by inflammatory group was detected for BDNF, VEGF-A or serotonin levels for any timepoint in the study (**Supplementary table 1**).

### Correlations between and within blood biomarkers and clinical scales

At each of the four major study timepoints, correlations were examined between blood biomarkers and study clinical scales (PCL-5, DASS-21 and WHO-5) to identify potential temporal relationships between these measures (**Figure 3, Supplementary Table 2**).

**Figure 3.**
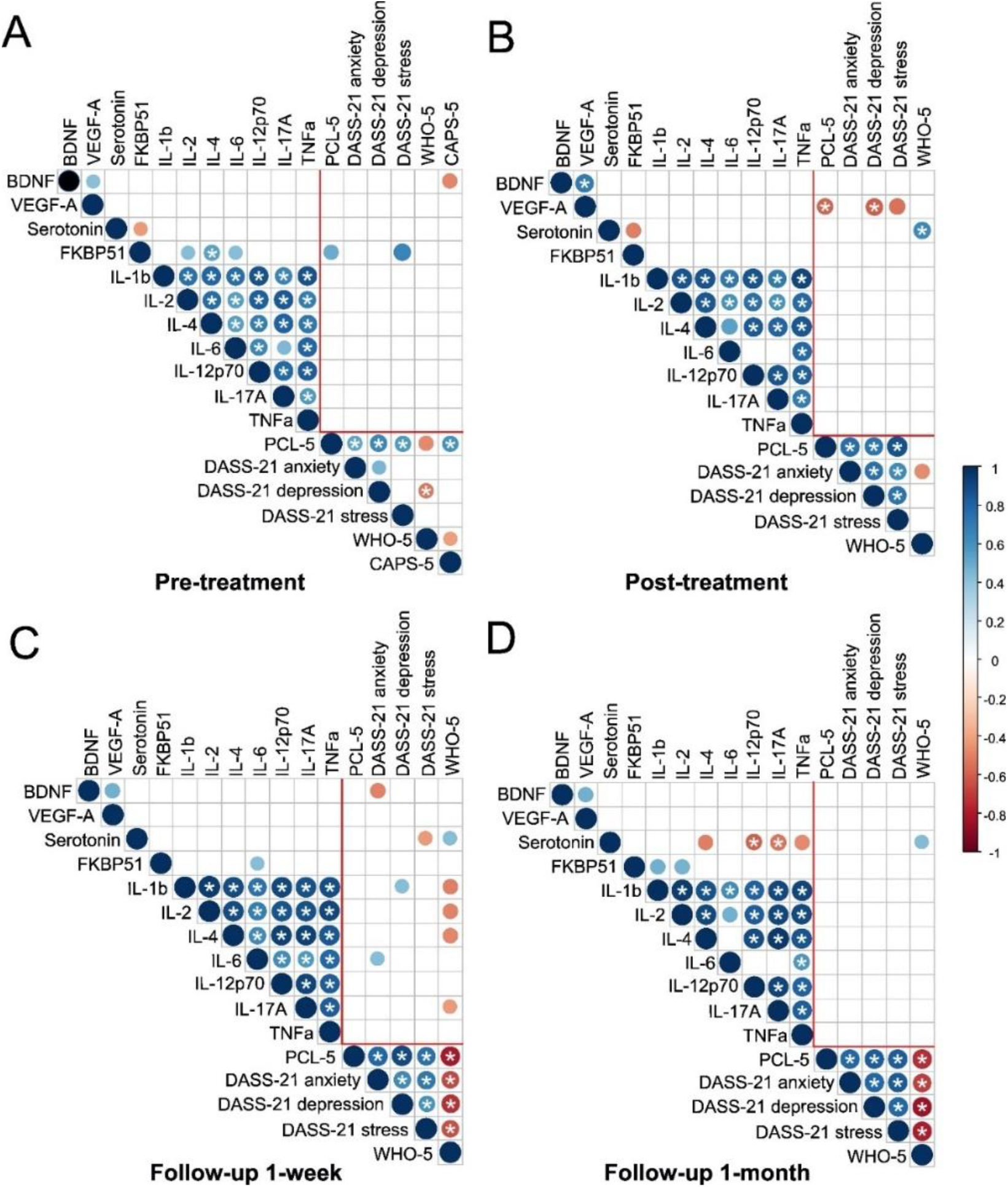
Spearman rank correlations between blood biomarkers and clinical scales at (A) pre-treatment, (B) post-treatment, (C) follow-up 1 week and (D) follow-up 1 month. Correlation coefficients are represented by the size and colour (key on right) of the circles. Only statistically significant correlations (p < 0.05) are shown. Correlations that remained significant after Benjamini-Hochberg (BH) false discovery rate adjustment for multiple comparisons contain asterisks. BDNF = Brain-derived neurotrophic factor, VEGF-A = Vascular endothelial growth factor A, FKBP51 = FK506-binding protein 51, PCL-5 = PTSD Checklist for DSM-5, DASS-21 = Depression Anxiety Stress Scales, WHO-5 = World Health Organisation Five Well-Being Index, CAPS-5 = Clinician-Administered PTSD Scale for DSM-5. Detailed correlation statistics are presented in **Supplementary Table 2**.

Notably, strong relationships were observed among immune cytokines (IL-1β, IL-2, IL-4, IL-6, IL-12p70, IL-17A and TNFα) and clinical scales (PCL-5, DASS-21 subscales and WHO-5) across study timepoints (**Figure 3**). Immune biomarker comparisons revealed only IL-6 diverged from the overall pattern of strong positive correlation between the cytokines at all the study timepoints. In terms of clinical scales, PCL-5 scores strongly correlated with the other scales at all the timepoints (including CAPS-5 scores at pre-treatment (r_s_(23)=0.593, p=0.002, p-adjusted=0.026)). DASS-21 subscales for anxiety, depression and stress and WHO-5 showed strengthening correlation as participants completed the ketamine treatment and continued to follow-up assessments (**Figure 3**).

Examining the non-immune blood biomarkers, BDNF and VEGF-A levels appeared to only correlate to each other, with positive correlations detected at all timepoints throughout the study (**Figure 3**; pre-treatment: r_s_(23)=0.415, p=0.039, p-adjusted=0.149; post-treatment: r_s_(17)=0.691, p=0.001, p-adjusted=0.006; follow-up 1-week: r_s_(22)=0.478, p=0.018, p-adjusted=0.054; follow-up 1-month: r_s_(21)=0.472, p=0.023, p-adjusted=0.075)). Serotonin levels were negatively correlated with FKBP51 at pre- and post-treatment (pre-treatment: r_s_(23)=0.415, p=0.039, p-adjusted=0.149; post-treatment: r_s_(17)=0.691, p=0.001, p-adjusted=0.006) and a subset of cytokines (IL-4, IL-12p70, IL-17A and TNFα) at follow-up 1-month (**Figure 3**). Finally, in addition to a relationship with serotonin, FKBP51 also exhibited sporadic correlations with cytokines IL-1β, IL-2, IL-4 and IL-6 at various timepoints across the trial (**Figure 3, Supplementary Table 2**).

Examining the relationships between blood biomarkers and clinical scales, several noteworthy correlations emerged. In relation to PTSD symptoms, only BDNF levels correlated negatively with CAPS-5 scores at pre-treatment (r_s_(23)=-0.487, p=0.014, p-adjusted=0.080) (**Figure 3A**), while FKBP51 levels correlated positively with PCL-5 at pre-treatment (r_s_(21)=0.497, p=0.016, p-adjusted=0.072) and VEGF-A levels correlated negatively with PCL-5 at post-treatment (r_s_(16)=-0.509, p=0.011, p-adjusted=0.039) (**Figure 3A & 3B**). Next, serotonin levels were consistently positively correlated with WHO-5 scores after ketamine treatment (post-treatment: r_s_(16)=0.632, p=0.005, p-adjusted=0.022; follow-up 1-week: r_s_(20)=0.425, p=0.48, p-adjusted=0.111; follow-up 1-month: r_s_(19)=0.440, p=0.046, p-adjusted=0.138) (**Figure 3, Supplementary Table 2**).

## Discussion

The present study represents the first comprehensive analysis of blood biomarkers to explore biological changes and relationships associated with ketamine treatment for PTSD. By investigating these mechanisms, our work takes an important step towards addressing the significant gap in understanding of how ketamine facilitates PTSD symptom improvement. We have identified key, albeit small, changes in BDNF and VEGF-A levels following treatment, along with significant associations between blood biomarkers and PTSD clinical measures.

BDNF has been extensively investigated across a range of mental health conditions, including PTSD. It assumes a crucial role in neuronal survival, growth and plasticity and, as such, is essential for learning and memory development^43^. Previous studies by our team (as well as many others) have reported relationships between circulating BDNF and PTSD^38, 44^. These relationships include correlations between BDNF levels and PTSD symptom severity^38, 44^, as well as differences in BDNF levels between cohorts with and without PTSD^21, 29^. Interestingly, while meta-analysis of the literature has found BDNF levels *lower* in cohorts with major depressive disorder (MDD), bipolar disorder, schizophrenia, panic disorder or obsessive-compulsive disorder, the same meta-analysis reported that BDNF levels are *higher* in cohorts with PTSD^21^. This observation suggests that PTSD may have distinct biological characteristics compared to the other psychiatric conditions, even though depression (as characterised by MDD) is often co-occurring. Moreover, it may be that BDNF might be considered a “goldilocks protein”, where levels that are either too low or too high contribute to the emergence of symptoms. Since elevated BDNF levels are often observed in PTSD, the small but significant decrease detected in our cohort following ketamine treatment (**Figure 2A**) may suggests a potential biological mechanism through which ketamine contributes to PTSD symptom improvement. It may be asserted that as PTSD symptoms ameliorate with ketamine, the brain’s requirement for elevated BDNF levels may diminish, resulting in a quantifiable decline. This supports the notion that BDNF increases in reaction to stress-induced neuronal damage but decreases when the stressor (i.e., PTSD symptoms) is mitigated^45^.

Interestingly, reducing elevated levels of BDNF may be a biomarker of general PTSD symptom improvement and not limited to just ketamine treatment. Zalta et al.^38^ recently reported the outcome of a 3-week cognitive processing therapy (CPT) treatment for military veterans with PTSD, showing that while BDNF levels at post-treatment study timepoints negatively correlated with PTSD symptoms, the change in each individual’s BDNF level from pre-treatment to post-treatment showed an overall significant decrease, similar to the findings in this study (**Figure 2A**). Similarly to our present study (**Table 1**), Zalta et al.^38^ also reported a wide variation in pre- and post-treatment BDNF levels among individual. This suggests that there may not be a universal “optimal” BDNF level, but rather that each individual has a unique baseline influenced by genetic or biological factors. Disruptions to this balance may only become apparent though broad comparisons between PTSD and control cohorts (as seen in previous literature^21, 29^) or through longitudinal individual BDNF analysis (as observed in this study and ^38^). Our previous work has also determined that the form of BDNF being measured (proBDNF verses mature BDNF verses total BDNF) is important in detecting correlations with PTSD symptom severity^44^, indicating that the relationship between BDNF and PTSD involves more dimensions than just a general amount in circulation.

The findings from the present study show a correlation between VEGF-A and BDNF levels (**Figure 3A-D**) and a parallel VEGF-A decrease after ketamine treatment (**Figure 1B**), supporting existing evidence that highlight the interdependence of BDNF and VEGF-A in mediating ketamine’s therapeutic effects. VEGF-A is best known for its roles in angiogenesis and blood vessel permeability but, like BDNF, it also has recognised neurotropic activity^46^. Both BDNF and VEGF-A have been linked genetically to antidepressant treatment responses in people^27^ and rodent studies have identified that the sustained antidepressant action of ketamine requires a reciprocal interdependence of both BDNF and VEGF-A on each other and the brain^28^. Rodent models of depression have found that ketamine rapidly increases BDNF and VEGF-A levels in the medial prefrontal cortex, which increases the number and function of synaptic spines, and in the hippocampus, which enhances neurogenesis and contributes to the antidepressant effects observed^47^. How these processes differ in PTSD (where our previous work has also detected elevated levels of VEGF-A in a PTSD cohort verses controls^29^) remains to be elucidated. Interestingly, at the post-treatment timepoint in this study, BDNF did not significantly correlate with any clinical scales, but VEGF-A levels significantly negatively correlated with PCL-5 and DASS depression and stress subscales and significantly positively correlated with BDNF at the same time (**Figure 3B**). Although small study sample sizes and individual variability in biomarkers can influence the detection of statistically significant relationships, the consistent associations observed between BDNF, VEGF-A and the clinical scales before and after treatment reinforce the notion that a biologically meaningful mechanism may be at play and that the present findings warrant further investigation.

The other group of significant biomarkers commonly investigated in relation to mental health (and PTSD) are the immune cytokines. A dysregulation of the immune system, commonly reported as a systemic, low-grade inflammation, has been observed in individuals with PTSD^22, 23^. In the present study, half of our trial participants appeared to have low-level inflammation, while the other half did not (**Figure 2**). The reason for this subgrouping was not apparent from the demographic information collected and did not appear to impact treatment response with ketamine. However, the variability in immune status within our PTSD cohort may help explain why other studies do not always detect significant immune differences related to PTSD status^48-50^. In the present study, there did not appear to be a detectable change in these circulating cytokine levels throughout the treatment and follow-up, but additional investigations with larger cohort sizes and broader, more sensitive panels are needed before any definitive statements can be made about immune biomarkers and changes related to ketamine treatment in PTSD.

Finally, despite their recognised roles in wellbeing and stress responses, research on serotonin and FKBP51 in the context of PTSD remains limited. Serotonin dysfunction has been linked to the pathophysiology of trauma-related symptoms associated with PTSD^51^, while abnormal increases in FKBP51 complexed with its glucocorticoid receptor appear to lead to glucocorticoid resistance, hyperarousal of the stress-response system, and symptoms of PTSD^25^. Previous research in mice has detected increased gene expression of FKBP51 in serotonergic neurons from the dorsal raphe after stress^52^, while human genotype-phenotype analysis has found FKBP51 single nucleotide polymorphisms influenced SSRI treatment outcomes in MDD^53^.

To our knowledge, this is the first study to report a significant relationship between circulating serotonin and FKBP51 levels, finding these biomarkers were negatively correlated during PTSD and immediately after treatment (**Figure 3A & 3B**). Notably, the correlation did not persist at the follow-up timepoints, prompting questions about how PTSD symptomology and its improvement may impact this relationship. Related to this was the fact that serotonin levels positively correlated with WHO-5 wellbeing scores, but only after ketamine treatment (**Figure 3B, 3C & 3D**). This corroborates other research that has reported whole blood serotonin levels can relate to wellbeing^54^, but that relationship was only seen as positively related to positive affect but not related to negative affect^55^. As such, the serotonin-wellbeing relationship was only detected once participants started feeling better. Alternatively, FKBP51 levels positively correlate with PCL-5 and DASS-21 stress scores at pre-treatment, suggesting its levels relate more strongly to the stress response linked to active PTSD.

There were limitations to this study that need to be acknowledged and considered when interpreting the results. The small sample size and incomplete sample sets for some participants limit the generalisability of the study findings. This should be considered, especially when certain changes or relationships were not detected (i.e. cytokine results). When relationships were detected, the small study numbers in combination with multiple comparisons caused several interesting correlations to lose statistical significance after p-value correction. To give the reader as much objective information as possible, both uncorrected and corrected p-vales were reported and relationships with supporting evidence in the literature were discussed. However, all the relationships detected in this study need replication in independent, larger cohorts. The samples for this analysis were also derived from an open-label (non-randomised, unblinded) clinical trial using oral ketamine for PTSD treatment and this may have introduced bias in the self-reported clinical scales. Furthermore, study participants were allowed to continue their existing medications during the ketamine treatment trial, thus the possibility of combined or synergic effects of ketamine with other medications cannot be ruled out. Despite these limitations, many statistically significant changes and relationships were still detectable within the study cohort, even after correcting for multiple comparisons, and findings were generally consistent or logical in the context of published literature.

In conclusion, this study reports the first comprehensive analysis of blood biomarker in participants with PTSD before and after ketamine treatment. Significant decreases in BDNF and VEGF-A levels were detected following treatment with ketamine, consistent with established knowledge that BDNF is typically elevated during PTSD and suggesting that BNDF may be a “goldilocks protein” that requires levels to be “just right” for the optimal mental health of individuals. This study adds new and significant information to our understanding of serotonin and FKBP51 in PTSD and symptom improvement. Finally, this study highlights the significant interpersonal variability in blood biomarker levels and reinforces the power of individual participant longitudinal analysis to understand important biological changes during illness and treatment.

## Data Availability

All data produced in the present study are available upon reasonable request to the authors.

## Acknowledgement

The authors express their sincere gratitude to the participants of the OKTOP study. The funding for this project was provided by the internal budget of the Thompson Institute. The authors declare that they have no conflict of interest.

## References

1. Benjet C, Bromet E, Karam EG, Kessler RC, McLaughlin KA, Ruscio AM et al. The epidemiology of traumatic event exposure worldwide: results from the World Mental Health Survey Consortium. Psychol Med 2016; 46(2): 327–343.

2. National Study of Mental Health and Wellbeing. https://www.abs.gov.au/statistics/health/mental-health/national-study-mental-health-and-wellbeing/latest-release, 2020-21, Accessed Date Accessed 2020-21 Accessed.

3. American Psychiatric Association. Diagnostic and statistical manual of mental disorders: DSM-5, vol. American Psychiatric Publishing, Inc.: Arlington, VA, US, 2013.

4. Green B. Post-traumatic stress disorder: New directions in pharmacotherapy. Advances in Psychiatric Treatment 2013; 19(3): 181–190.

5. Thomas E, Stein DJ. Novel pharmacological treatment strategies for posttraumatic stress disorder. Expert Rev Clin Pharmacol 2017; 10(2): 167–177.

6. López-Ojeda W, Hurley RA. Extended Reality Technologies: Expanding Therapeutic Approaches for PTSD. J Neuropsychiatry Clin Neurosci 2022; 34(1): A4–5.

7. Klaeth JR, Jensen AG, Auren TJB, Solem S. 12-month follow-up of intensive outpatient treatment for PTSD combining prolonged exposure therapy, EMDR and physical activity. BMC Psychiatry 2024; 24(1): 225.

8. Asnis GM, Kohn SR, Henderson M, Brown NL. SSRIs versus Non-SSRIs in Post-traumatic Stress Disorder. Drugs 2004; 64(4): 383–404.

9. Berger W, Mendlowicz MV, Marques-Portella C, Kinrys G, Fontenelle LF, Marmar CR et al. Pharmacologic alternatives to antidepressants in posttraumatic stress disorder: a systematic review. Prog Neuropsychopharmacol Biol Psychiatry 2009; 33(2): 169–180.

10. Nikolin S, Rodgers A, Schwaab A, Bahji A, Zarate C, Jr., Vazquez G et al. Ketamine for the treatment of major depression: a systematic review and meta-analysis. EClinicalMedicine 2023; 62: 102127.

11. Ragnhildstveit A, Roscoe J, Bass LC, Averill CL, Abdallah CG, Averill LA. The potential of ketamine for posttraumatic stress disorder: a review of clinical evidence. Ther Adv Psychopharmacol 2023; 13: 20451253231154125.

12. Quigley BL, Can AT, Dutton M, Gallay CC, Forsyth G, Jones M et al. Low dose oral ketamine treatment on post-traumatic stress disorder (PTSD) (OKTOP): An open-label pilot study. European Neuropsychopharmacology 2025; 93: 17–18.

13. Andrade C. Oral Ketamine for Depression, 1: Pharmacologic Considerations and Clinical Evidence. J Clin Psychiatry 2019; 80(2).

14. Can AT, Hermens DF, Dutton M, Gallay CC, Jensen E, Jones M et al. Low dose oral ketamine treatment in chronic suicidality: An open-label pilot study. Transl Psychiatry 2021; 11(1): 101.

15. Muhorakeye O, Biracyaza E. Exploring Barriers to Mental Health Services Utilization at Kabutare District Hospital of Rwanda: Perspectives From Patients. Front Psychol 2021; 12: 638377.

16. Berman RM, Cappiello A, Anand A, Oren DA, Heninger GR, Charney DS et al. Antidepressant effects of ketamine in depressed patients. Biol Psychiatry 2000; 47(4): 351–354.

17. Johnston JN, Zarate CA, Kvarta MD. Esketamine in depression: putative biomarkers from clinical research. European Archives of Psychiatry and Clinical Neuroscience 2024.

18. Kumar R, Nunez NA, Joshi N, Joseph B, Verde A, Seshadri A et al. Metabolomic biomarkers for (R, S)-ketamine and (S)-ketamine in treatment-resistant depression and healthy controls: A systematic review. Bipolar Disord 2024; 26(4): 321–330.

19. Pradhan B, Mitrev L, Moaddell R, Wainer IW. d-Serine is a potential biomarker for clinical response in treatment of post-traumatic stress disorder using (R,S)-ketamine infusion and TIMBER psychotherapy: A pilot study. Biochim Biophys Acta Proteins Proteom 2018; 1866(7): 831–839.

20. Michopoulos V, Norrholm SD, Jovanovic T. Diagnostic biomarkers for posttraumatic stress disorder: Promising horizons from translational neuroscience research. Biol Psychiatry 2015; 78(5): 344–353.

21. Zou Y, Zhang Y, Tu M, Ye Y, Li M, Ran R et al. Brain-derived neurotrophic factor levels across psychiatric disorders: A systemic review and network meta-analysis. Progress in Neuro-Psychopharmacology and Biological Psychiatry 2024; 131: 110954.

22. Hori H, Kim Y. Inflammation and post-traumatic stress disorder. Psychiatry Clin Neurosci 2019; 73(4): 143–153.

23. Baker DG, Nievergelt CM, O’Connor DT. Biomarkers of PTSD: neuropeptides and immune signaling. Neuropharmacology 2012; 62(2): 663–673.

24. Oglodek EA. Changes in the Serum Concentration Levels of Serotonin, Tryptophan and Cortisol among Stress-Resilient and Stress-Susceptible Individuals after Experiencing Traumatic Stress. Int J Environ Res Public Health 2022; 19(24).

25. Li H, Su P, Lai TK, Jiang A, Liu J, Zhai D et al. The glucocorticoid receptor-FKBP51 complex contributes to fear conditioning and posttraumatic stress disorder. J Clin Invest 2020; 130(2): 877–889.

26. Mehta D, Gonik M, Klengel T, Rex-Haffner M, Menke A, Rubel J et al. Using polymorphisms in FKBP5 to define biologically distinct subtypes of posttraumatic stress disorder: evidence from endocrine and gene expression studies. Arch Gen Psychiatry 2011; 68(9): 901–910.

27. Kao CF, Liu YL, Yu YW, Yang AC, Lin E, Kuo PH et al. Gene-based analysis of genes related to neurotrophic pathway suggests association of BDNF and VEGFA with antidepressant treatment-response in depressed patients. Sci Rep 2018; 8(1): 6983.

28. Deyama S, Bang E, Kato T, Li XY, Duman RS. Neurotrophic and antidepressant actions of brain-derived neurotrophic factor require vascular endothelial growth factor. Biol Psychiatry 2019; 86(2): 143–152.

29. Quigley BL, Wellington N, Levenstein JM, Dutton M, Boucas AP, Forsyth G et al. Circulating biomarkers and neuroanatomical brain structures differ in older adults with and without post-traumatic stress disorder (PTSD). Sci Rep 2025; 15: 7176.

30. Weathers FW, Bovin MJ, Lee DJ, Sloan DM, Schnurr PP, Kaloupek DG et al. The Clinician-Administered PTSD Scale for DSM-5 (CAPS-5): Development and initial psychometric evaluation in military veterans. Psychol Assess 2018; 30(3): 383–395.

31. Blevins CA, Weathers FW, Davis MT, Witte TK, Domino JL. The Posttraumatic Stress Disorder checklist for DSM-5 (PCL-5): Development and initial psychometric evaluation. J Trauma Stress 2015; 28(6): 489–498.

32. Lovibond SH, Lovibond PF. Manual for the Depression Anxiety Stress Scales (2nd Ed.). Psychology Foundation: Sydney, 1995.

33. Bech P, Olsen LR, Kjoller M, Rasmussen NK. Measuring well-being rather than the absence of distress symptoms: a comparison of the SF-36 Mental Health subscale and the WHO-Five Well-Being Scale. Int J Methods Psychiatr Res 2003; 12(2): 85–91.

34. R Core Team. R: A Language and Environment for Statistical Computing, R Foundation for Statistical Computing. Vienna, Austria, https://www.R-project.org/, 2024.

35. Bates D, Mächler M, Bolker B, Walker S. Fitting Linear Mixed-Effects Models Using lme4. Journal of Statistical Software 2015; 67(1): 1 – 48.

36. Kuznetsova A, Brockhoff PB, Christensen RHB. lmerTest Package: Tests in Linear Mixed Effects Models. Journal of Statistical Software 2017; 82(13): 1 – 26.

37. Wickham H. Ggplot2: Elegant graphics for data analysis (2nd ed.). Springer International Publishing: Cham, Switzerland, 2016.

38. Zalta AK, Voigt RM, Stevens SK, Held P, Raeisi S, Boley RA et al. Brain derived neurotrophic factor and treatment outcomes among veterans attending an intensive treatment program for posttraumatic stress disorder. J Psychiatr Res 2024; 173: 1–5.

39. gplots: Various R Programming Tools for Plotting Data. 2015.

40. Benjamini Y, Hochberg Y. Controlling the False Discovery Rate: A Practical and Powerful Approach to Multiple Testing. Journal of the Royal Statistical Society: Series B (Methodological) 1995; 57: 289–300.

41. Harrell Jr F. Hmisc: Harrell Miscellaneous. R package version 5.2-2, https://github.com/harrelfe/hmisc. 2025.

42. Wei SV. R package ‘corrplot’: Visualization of a Correlation Matrix. (Version 0.95), https://github.com/taiyun/corrplot. 2024.

43. Bathina S, Das UN. Brain-derived neurotrophic factor and its clinical implications. Arch Med Sci 2015; 11(6): 1164–1178.

44. Quigley BL, Wellington N, Dutton M, Bouças AP, Forsyth G, Gallay CC et al. Mature brain-derived neurotrophic factor (BDNF) levels in serum correlate with symptom severity in post-traumatic stress disorder (PTSD). Journal of Affective Disorders Reports 2025; 19: 100854.

45. Guo W, Pang K, Chen Y, Wang S, Li H, Xu Y et al. TrkB agonistic antibodies superior to BDNF: Utility in treating motoneuron degeneration. Neurobiology of Disease 2019; 132: 104590.

46. Sondell M, Lundborg G, Kanje M. Vascular endothelial growth factor has neurotrophic activity and stimulates axonal outgrowth, enhancing cell survival and Schwann cell proliferation in the peripheral nervous system. J Neurosci 1999; 19(14): 5731–5740.

47. Deyama S, Duman RS. Neurotrophic mechanisms underlying the rapid and sustained antidepressant actions of ketamine. Pharmacol Biochem Behav 2020; 188: 172837.

48. Passos IC, Vasconcelos-Moreno MP, Costa LG, Kunz M, Brietzke E, Quevedo J et al. Inflammatory markers in post-traumatic stress disorder: a systematic review, meta-analysis, and meta-regression. Lancet Psychiatry 2015; 2(11): 1002–1012.

49. Renna ME, O’Toole MS, Spaeth PE, Lekander M, Mennin DS. The association between anxiety, traumatic stress, and obsessive-compulsive disorders and chronic inflammation: A systematic review and meta-analysis. Depress Anxiety 2018; 35(11): 1081–1094.

50. Yang JJ, Jiang W. Immune biomarkers alterations in post-traumatic stress disorder: A systematic review and meta-analysis. J Affect Disord 2020; 268: 39–46.

51. Davis LL, Suris A, Lambert MT, Heimberg C, Petty F. Post-traumatic stress disorder and serotonin: new directions for research and treatment. J Psychiatry Neurosci 1997; 22(5): 318–326.

52. Lesiak AJ, Coffey K, Cohen JH, Liang KJ, Chavkin C, Neumaier JF. Sequencing the serotonergic neuron translatome reveals a new role for Fkbp5 in stress. Mol Psychiatry 2021; 26(9): 4742–4753.

53. Ellsworth KA, Moon I, Eckloff BW, Fridley BL, Jenkins GD, Batzler A et al. FKBP5 genetic variation: association with selective serotonin reuptake inhibitor treatment outcomes in major depressive disorder. Pharmacogenet Genomics 2013; 23(3): 156–166.

54. de Vries LP, van de Weijer MP, Bartels M. The human physiology of well-being: A systematic review on the association between neurotransmitters, hormones, inflammatory markers, the microbiome and well-being. Neuroscience & Biobehavioral Reviews 2022; 139: 104733.

55. Williams E, Stewart-Knox B, Helander A, McConville C, Bradbury I, Rowland I. Associations between whole-blood serotonin and subjective mood in healthy male volunteers. Biological Psychology 2006; 71(2): 171–174.

